# Identifying factors related to nondisclosure during clinical supervision among Chinese supervisees: Study protocol

**DOI:** 10.1101/2024.02.02.24302184

**Authors:** Alberto Stefana, Eric A. Youngstrom

## Abstract

Psychoanalysis is gaining widespread popularity as a therapeutic model in China. However, the absence of local accredited psychoanalytic training institutes necessitates that Chinese trainees and clinicians often receive supervision from European or American supervisors through online platforms. A key component of effective supervision is the willingness of the supervisees to disclose. This openness not only solidifies the supervisory alliance but also improves the supervisee’s self-awareness, self-efficacy, knowledge, and therapeutic skills. Emerging research highlights the significant role of cultural factors in shaping supervisory relationships. Our study aims to: (a) compare the disclosure and nondisclosure rates in clinical supervision between Chinese supervisees and their counterparts from Western cultures, and (b) examine the association between these behaviors and variables related to the therapeutic process among Chinese supervisees. We will employ a cross-sectional design, targeting Chinese psychoanalytic clinicians, both trainees and licensed professionals, engaged in individual clinical supervision. Recruitment will be primarily through the China American Psychoanalytic Alliance mailing list, complemented by snowball sampling. Data will be collected through an online survey hosted on the REDCap platform, focusing on the primary clinical supervisor of the participants and their supervisory experience. This research aims to provide insights that could enable supervisors of Chinese trainees to better understand and adapt to the relational dynamics in supervisory contexts. Additionally, it will lay the foundation for more detailed qualitative investigations into the experiences of Chinese psychoanalytic supervisees, potentially informing future supervisory practices and training methodologies.

---

Supervision is the centerpiece of psychoanalytic training [1–3] and serves the dual function of making supervisees become competent clinicians [4] while improving patients’ outcomes [5].

Supervisees’ self-disclosure—which is a realistic description of the process and outcomes of their patients’ therapy, as well as an open sharing of their in- and out-session feelings, thoughts, and behavior toward their patients and supervisor—is a central asset for effective supervision.

Disclosure can strengthen the supervisory alliance and make the supervisee feel security in supervision [6,7] and increase self-awareness, self-efficacy, knowledge, and therapeutic skills [4,8].

Despite the above, nondisclosure of important clinical information appears to be a frequent and normative aspect of supervision that hinders its effectiveness and contributes to the loss of potential learning experiences [9–12]. Although supervisees, especially when at the beginning of their training, do not always know what kinds of personal and clinical material are most important for supporting and enhancing the supervision process, empirical evidence indicates that many trainees deliberately withhold information that they know is relevant for supervision (e.g., [13]), usually for fear of judgment or negative evaluation [14]. This is especially likely when supervisees felt negatively about their supervisor or the supervision experience (e.g., [15,16]). The decision to avoid bringing up difficulties with patients or dissatisfaction with the supervision pathway is often due to the evaluative nature of supervision [5].

The consequences of withholding relevant clinical-related information are problematic, especially when the supervisor does not have access to audio- or video-recordings of the therapy sessions [9,17]—probably the typical condition in psychoanalytic training. Furthermore, nondisclosure of clinical material limits the ability of the supervisor to correctly evaluate the supervisee’s activity and guide them through their work with patients [5]. This, in extreme cases, may mean having the supervisor unaware of situations that could result in serious ethical violations (e.g., sexual misconduct) or negative consequences for the health and well-being of the patient.

Supervision-related nondisclosure is troubling too. When a supervisee withholds from revealing the feeling that supervision is not helpful or even harmful [18,19], the supervisor is unlikely to know about the necessity of adjusting their manner of conducting supervision in order to become more responsive to supervisee’s training needs [20,21]. Understandably, trainees often are reluctant to express a negative perception of the supervision process or the supervisor themselves. Psychoanalytic training occurs in a work environment, and it requires courage for subordinates to confront a supervisor. Concerns about appearing disrespectful or being negatively evaluated come into play [22]. In psychoanalytic training, for example, a trainee who challenges a supervisor may be seen as someone having problems with authority.

Psychoanalysis is currently one of the most popular and influential therapeutic models in China. It was present in Chinese psychiatry since the early 20th century [23] and was among the therapeutic approaches that entered China with the opening of doors to Western intellectual products in the 1980s [24]. As China has transported psychoanalytic models of psychotherapy from the West [25–27], they started a complex dialogic process as Chinese clinicians have been embracing, selecting, and reworking strands of psychological knowledge they have been importing [28]. Furthermore, due to the lack of on-site accredited psychoanalytic training institutes, it has become routine for Chinese trainees to be analyzed and supervised by European or United States psychoanalysts and psychoanalytic psychotherapists via Zoom or Skype [29]. The China American Psychoanalytic Alliance (CAPA) is an association born to “develop and promote mental health services in China by training Chinese mental health professionals as psychodynamic psychotherapists and providing them with psychoanalytic and psychotherapeutic treatment” (CAPA website). Since the start of the new millennium, CAPA members, who are predominantly American (but also Australian, Canadian, European, Israeli, and South and Central American), have trained more than 2,000 volunteers and mental health professionals. Growing evidence shows that distance psychoanalytic training works [23,30,31]; however, certain cultural specificities [32–35] raise questions about the dynamics in play within the supervision of Chinese trainees. These dynamics may be unknown or difficult to recognize, especially by non-Chinese supervisors.

Despite the popularity of psychoanalysis and the key role of supervision in training, little work has previously focused on this topic with a focus on Chinese trainees. However, there is a network of research that speaks to the importance of the topic and informs hypotheses. Literature on Chinese culture and communication shows that: speaking the same dialect is associated with a high quality of the subordinate–supervisor relationship [36]; leaders/supervisors have both in- and out-group relationships with their employees/supervisees [37,38]; social encounters or interactions foster group bonding and promotes social harmony [39]. Therefore, Chinese people usually have more opportunities to communicate with supervisors in non-work contexts, and speaking the same dialect can be a facilitating factor for subordinates to build a closer relationship with the supervisor [40]. At work, in line with a tradition of hierarchy that can be traced back to Confucian tradition and vertical cultural values system, Chinese experience a strong obligation to obey their superiors [41,42]. These observations suggest that Chinese employees/supervisees’ beliefs in mutual obligations between themselves and the employer/supervisor are characterized by asymmetric power [42]. Added to and incorporated into all this is the issue of differences in nonverbal communication, in fact, there are several nonverbal factors used in daily life that may cause misunderstanding in Chinese and Western cross-cultural communication.

A recent descriptive study comparing clinical supervision practices in seven countries (including China) found cultural differences in terms of forms of communication, power, manifestations of respect, supervision hierarchy, and cultural relevance of evaluation, feedback, regulation, and gatekeeping [43]. These findings align with previous observations of the supervisory relationship processes in countries influenced by Confucian traditions, which show that the supervisor-supervisee system is influenced by filial piety and attention to hierarchy as well as the importance of harmony and preserving reputation [44,45].

Given the large and rapidly growing demand for this type of training, it is crucial to study and optimize the supervising relationship using relevant data. However, the scope of the empirical literature is largely limited to Western supervision practices, and most research primarily investigates supervisee nondisclosure, often as a unidimensional construct and rarely as a two-dimensional multivariate composite, using measures that lack adequate psychometric properties.

## Study objectives

In light of the above, the main objectives of this study are to explore:

1. Similarities and differences between Chinese supervisees and Western culture supervisees in terms of disclosure and nondisclosure rates in individual clinical supervision;
2. whether Chinese supervisees’ disclosure and nondisclosure behaviors are associated with three important process variables:
  a. the quality of the supervisory relationship (including alliance),
  b. the supervisor’s style and relational approach, and
  c. the satisfaction with the clinical supervision.
  Achieving these objectives will:

i. improve measurement of supervision process,
ii. identify “pain points” in supervision with Chinese supervisees, and
iii. provide tools for measuring improvements in supervision.

## Hypotheses

From a comparative perspective between samples (prior results from, e.g., [22,46]), we hypothesize that Chinese supervisees will exhibit lower average disclosure rates compared to their counterparts in the United States and other Western countries. This prediction is premised on the differing cultural traditions of hierarchy. We hypothesize that a higher extent of both clinically related and supervision-related nondisclosures and lower willingness to disclose in supervision will be reported by supervisees who report higher levels of anxiety and lower levels of self-esteem, experience a weaker supervisory relationship, and perceive their supervisors as low in attractiveness and interpersonal sensitiveness. Lastly, we hypothesize that less satisfied supervisees will have more nondisclosures and negative reasons for not disclosing, both characterized by a negative tone. Moreover, we hypothesize that after controlling for other predictors, each variable will individually predict nondisclosures, demonstrating incremental validity. Additionally, we propose that the three process variables will jointly contribute to predicting nondisclosures.

## Methods

### Study design

This is an observational study with a cross-sectional design. Participants are Chinese psychoanalytic psychotherapy clinicians, either in training or licensed/qualified, under individual clinical supervision.

### Measures

This study will use a comprehensive set of self-report scales to collect extensive data on the supervision process and outcome. The data will cover six main domains. The personal and professional characteristics domain captures the supervisee’s demographic, professional, and supervision characteristics. The supervisee functioning domain focuses on the supervisee’s levels of state anxiety regarding the attendance of supervision sessions and his/her energetic and effective connection with the clinical work. The supervisor attitude and style domain evaluates the supervisor’s style, cultural humility, and focus on relational behavior. The relationship and collaboration domain assesses the quality of specific elements of the supervisory relationship between the supervisee and the clinical supervisor. The disclosure domain investigates the supervisee’s willingness to disclose and intentionality to withdraw information in clinical supervision. Lastly, the impact and outcome domain assesses the supervisee’s level of satisfaction with their clinical supervision, as well as the perceived influence of the supervision on both him/herself and the patients he/she serves. All domains are assessed from the supervisee’s perspective.

The survey contains 200 items and can be completed in approximately 25 minutes.

#### Personal and professional characteristics domain

A *Demographic and professional data form* will collect information such as respondents’ age, biological sex, gender, level of training, months of psychotherapy experience, the average number of patients seen per week, date supervision started, hours of supervision per week, supervisor’s sex, and supervisor’s race. The data form consists of 22 items.

#### Supervisee functioning domain

The *Anticipatory Supervisee Anxiety Scale* (ASAS) [47] is a self-report measure comprising 28 items that evaluate the affective and cognitive aspects of supervisee anxiety in anticipation of supervision sessions. Respondents rate items on a 9-point Likert scale ranging from “Not at all” (1) to “Completely true” (9), with the ASAS demonstrating excellent reliability (Cronbach’s alpha = .97; Tosado, 2004).

The *Utrecht Work Engagement Scale* (UWES) [48], in its ultra-short, 3-item format, assesses vigor, dedication, and absorption, using a 6-point Likert scale from “Never” (1) to “Always/Every day” (6). The scale showed good internal consistency with Cronbach’s alpha values from .83 to .91. The UWES has also proven effective in cross-cultural contexts, including Chinese work environments [49].

#### Supervisor attitude and style domain

The *Supervisory Styles Inventory* (SSI) [50] assesses supervisees’ perceptions of their supervisors’ styles through a 33-item self-report scale, including attractive (e.g., flexible and supportive), interpersonally sensitive, and task-oriented subscales. Ratings range from “Not very” (1) to “Very” (7), with eight filler items included. The SSI exhibits high internal consistency, with Cronbach’s alpha ranging from .89 to .91 for the full scale and .89-.93, .88, and .84-.85 for the attractive, interpersonally sensitive, and task-oriented subscales, respectively.

The *Relational Behavior Scale* (RBS) [51] is an 11-item self-report instrument designed to gauge supervisees’ perceptions of their supervisors’ relational behaviors in the most recent session, such as exploration of feelings and focus on countertransference. Responses are given on a Likert-type scale from “Not at all” (1) to “Very much” (5). The RBS demonstrated good reliability with a Cronbach alpha of .85.

The Cultural Humility Scale (CHS) [52] measures the perceived extent of supervisors’ respect for others’ cultural orientations and values, using a 12-item self-report format. Rated on a 5-point Likert scale from “Strongly disagree” (1) to “Strongly agree” (5), including five reversed-scored items, the CHS has shown strong reliability, with Cronbach’s alpha coefficients between .86 and .93 for the total scale, .90 for the positive subscale, and .88-.90 for the positive items.

#### Relationship and collaboration domain

The *Supervisory Working Alliance–Supervisee version* (SWA-S) [53] is a tool designed to capture supervisees’ perceptions of the quality of the supervisory working alliance. With 19 items rated on a 7-point Likert scale from “Almost never” (1) to “Almost always” (7), the SWA-S includes rapport (12 items) and client focus (6 items) subscales. It has shown excellent internal consistency, with Cronbach’s alpha coefficients ranging from .90 to .97 for rapport, .87 to .94 for client focus, and .95 to .98 for the total scale, in various studies [16,54–56].

The *Collaborative Behavior Supervision Scale* (CBSS) [57] assesses collaborative clinical supervision from the point of view of the supervisee through a concise four-item self-report scale. Collaboration, defined as the mutual agreement and cooperative effort in clinical supervision, is rated on a 5-point Likert scale from “Never” (1) to “Always” (5), with the CBSS showing a high Cronbach’s alpha of .93, indicating robust reliability.

The *Short Supervisory Relationship Questionnaire* (S-SRQ) [58] evaluates the supervisory relationship within clinical supervision settings. This 18-item self-report scale rates responses from “Strongly Disagree” (1) to “Strongly Agree” (7) and encompasses three subscales: safe base (9 items), reflective education (5 items), and structure (4 items). The overall internal reliability of the S-SRQ is exemplary, with a Cronbach alpha of .96, and its subscales show alphas of .97 for safe base, .89 for reflective education, and .88 for structure, supporting its use in the assessment of various dimensions of supervisory relationships.

The *Supervisee in-Session Affective Reactions Scale* (SiSARS), adapted from the *in-Session Patient Affective Reactions Questionnaire* (SPARQ) [59]. The SPARQ is an eight-item self-report tool designed to gauge a patient’s emotional responses towards their therapist during a psychotherapy session. This measure consists of two separate, four-item scales yielding two distinct scores that are not intended to be combined. The Positive Affect scale measures the patient’s sense of a safe and comforting therapeutic bond, while the Negative Affect scale encompasses feelings of embarrassment, timidity, reluctance to speak freely, concerns over insufficient support, and a sense of inadequacy related to seeking assistance from the therapist. Each item is assessed using a 5-point Likert scale, ranging from 0 (“Not at all true”) to 4 (“Very true”). For this study, the SiSARS was adjusted to apply to clinical supervision contexts, with “therapist” being substituted with “supervisor” in the instructions and item descriptions.

#### Disclosure domain

The *Supervisee Nondisclosure Scale* (SNDS) [60,61] measures the frequency of intentional nondisclosure of relevant information by supervisees to their supervisors. The scale consists of 30 items rated on a 3-point Likert scale with options “Fully disclosed” (1), “Somewhat disclosed” (2), and “Decided to not disclose” (3), where higher scores denote greater nondisclosure. It features two subscales: supervision-related nondisclosure (4 items) and clinically-related nondisclosure (7 items), with reported marginal reliabilities of .83 and .77, respectively.

The *Trainee Disclosure Scale* (TDS) [62] evaluates the propensity of supervisees to openness in clinical supervision through a 13-item self-report measure. Items are rated on a 5-point Likert scale from “Not at all likely” (1) to “Very likely” (5). Higher scores on the TDS reflect greater willingness to disclose, and it has demonstrated high internal consistency with Cronbach alpha coefficients equal to or exceeding .86, as evidenced in multiple studies [13,62,63].

#### Impact and outcome domain

The *Supervisory Satisfaction Questionnaire–Supervisees form* (SSQ) [10] quantifies supervisees’ satisfaction with their clinical supervision. This 8-item scale employs a 4-point Likert-type scale to gauge responses, revealing a unidimensional factor structure. The SSQ has shown strong internal consistency with the Cronbach alpha ranging from .84 to .93.

The *Supervision Outcome Scale* (SOS) [64]is designed to evaluate the effectiveness of clinical supervision from the supervisee’s perspective, encompassing effects on the supervisee and their patients. This 7-item scale is divided into two subscales assessing clinical competence (4 items) and multicultural competence (3 items), with each item rated on a 5-point Likert scale from “Not helpful at all” (1) to “Extremely helpful” (5). The SOS has shown excellent reliability with a Cronbach alpha of .90 for the overall scale, and subscale alphas of .86 for clinical competence and .94 for multicultural competence.

## Statistical analysis

We will employ descriptive statistics to examine demographic details and the distribution of scores across various measures. For ease of interpretation, we will use independent sample *t*-tests to contrast the mean scores on each measure for Chinese supervisees with those reported for non-Chinese supervisees in the existing literature, and calculate Cohen’s *d* as an effect size for the differences. We intend to examine the content of and reasons for Chinese supervisees’ disclosure *vs* nondisclosure in supervision. Variations in (non)disclosure patterns, as linked with demographic, professional, and supervision characteristics, will be assessed in line with our first objective. To evaluate the potential impact of the sequence of sections of the survey on the variables studied, a series of one-way MANOVAs will be conducted. If order effects prove significant, the order will be included as a covariate in subsequent analyses. We will determine the correlations between demographic characteristics and both disclosure and nondisclosure behaviors. Finally, multivariate multiple regression analyses will be undertaken to evaluate the combined and individual contributions of our predictors (i.e., supervisees’ perceptions of the supervisory relationship, their supervisor’s style and relational approach, and perceived satisfaction with supervision) to the dependent variables (i.e., disclosure and nondisclosure). This step corresponds to our second objective.

### Eligibility criteria

Participants are eligible for the study if they are Chinese psychoanalysts or psychoanalytic psychotherapists (either in training or licensed/qualified), are 18 years or older, are fluent in English, and currently are under individual clinical supervision.

### Recruitment

Participants will be psychoanalysis and psychoanalytic psychotherapy supervisees recruited through the mailing lists of the China American Psychoanalytic Alliance (CAPA) and other psychological or psychoanalytic associations. Potential participants will receive an email containing a link to the online survey website where they can access the questionnaire. Potential participants also will receive three follow-up notifications to remind them about the questionnaire. Snowball sampling will be also used in that participants are asked to forward the study link to other Chinese psychoanalytic clinicians of their acquaintance.

### Data collection process

Participants will be involved in a one-time anonymous survey conducted using the Research Electronic Data Capture (REDCap) software platform. They will be required to complete the full suite of measures provided. Those with more than one supervisor will be instructed to select their main individual clinical supervisor and to focus on the supervisory relationship and work conducted with this person when responding to the measures.

### Ethical considerations

This study has been granted an exemption by the Institutional Review Board (IRB) at the University of North Carolina at Chapel Hill (Study #: 23-2796).

As the initial screen of the online survey, participants will be presented with a comprehensive digital consent document. This document will detail the study’s objectives, underlying reasoning, and methodology, in addition to providing the contact information for both the principal investigator and the Institutional Review Board (IRB). The consent document will clarify to potential participants that in order to protect their identities, only relevant and minimally necessary information will be collected. Moreover, it will ensure that any published results will focus on groups, not individual participants. At the bottom of this participant information sheet, there will be a declaration of consent. Here, potential participants must choose between participating in the study and declining. Those who do not consent will be redirected to a final page. The document will clearly state that all participants retain the right to withdraw from the study at any time, without having to provide an explanation.

### Dissemination policy

The findings of this research will initially be made public in the form of preprints, followed by dissemination in peer-reviewed journals and through presentations at scientific conferences. A dedicated repository on Open Science Framework has been established to house at the link https://osf.io/tfx8b the study’s scales (only those not protected by copyright), dataset, presentations, and preliminary preprint manuscripts. Additionally, the insights obtained from the study could be disseminated to relevant mental health organizations, thus informing future research and potentially improving psychotherapeutic practices.

## Discussion

Insights from this research may offer supervisors of Chinese supervisees a deeper understanding of the relational process in supervisory situations. This could potentially enable them to adjust their technical frames when necessary. Given that supervisees’ experiences during supervision can significantly impact the supervisory relationship and subsequent patient treatment, it is important that supervisors incorporate knowledge of their Chinese supervisees’ experiences into their clinical supervision strategy. Furthermore, this study will likely guide future qualitative research by providing a roadmap for an in-depth exploration of Chinese supervisees’ experiences.

## Data Availability

Once the study is completed, data can be obtained from the OSF.io website at this URL: https://osf.io/tfx8b.

## Acknowledgment

None.

